# Seroprevalence of chikungunya and o’nyong-nyong viruses in Senegal, West Africa

**DOI:** 10.1101/2024.09.06.24313176

**Authors:** Prince Baffour Tonto, Mouhamad Sy, Ibrahima Mbaye Ndiaye, Mariama Toure, Amy Gaye, Mariama Aidara, Amadou Moctar Mbaye, Abdoulaye Kane Dia, Mamadou Alpha Diallo, Jules Francois Gomis, Mamadou Samba Yade, Younous Diedhiou, Baba Dieye, Khadim Diongue, Mame Cheikh Seck, Aida S. Badiane, Bobby Brooke Herrera, Daouda Ndiaye

## Abstract

**Background:** Arthritogenic alphaviruses such as chikungunya (CHIKV) and o’nyong-nyong (ONNV) viruses have shown capacity to cause widespread epidemics, with recurrent and sporadic outbreaks occurring throughout sub-Saharan Africa.

**Methods:** We analyzed the seroprevalence for CHIKV and ONNV in 470 non-febrile subjects from three regions in Senegal (Sindia, 2018; Thies, 2018; and Kedougou, 2022/2023) using retrospective samples. We assessed the presence of anti-CHIKV IgG and neutralizing antibody titers against CHIKV and ONNV via enzyme-linked immunosorbent assay (ELISA) and microneutralization tests, respectively, and determined risk factors of CHIKV and ONNV exposure by binary logistic regression.

**Results:** The overall alphavirus seroprevalence based on an anti-CHIKV viral like particle (VLP) IgG ELISA was 38.5%, with rates varying geographically: Kedougou (48.6%), Thies (31.9%), and Sindia (14.9%). Neutralizing antibody titers revealed CHIKV and ONNV seroprevalence rates of 7.4% and 9.8%, respectively, with significant variations by region and age group. Cross-reactivity analysis showed that 82.9% of CHIKV cases exhibited a neutralizing response to ONNV, while 71.7% of ONNV cases cross-neutralized CHIKV. Residents of Thies had significantly higher odds of CHIKV infection (aOR, 3.147; 95% CI: 1.164-8.510) while ONNV infection was more likely in Kedougou (aOR, 3.888; 95% CI: 1.319-11.466). Furthermore, older age (> 40 years) was a significant risk factor both CHIKV (aOR, 2.094; 95% CI: 0.846-5.185) and ONNV infection (aOR, 2.745; 95% CI: 1.212-6.216).

**Conclusions:** Our study confirms the co-circulation of CHIKV and ONNV in Senegal, highlighting their geographic and demographic distribution. These findings underscore the need for continued surveillance, alphavirus testing, and tailored public health strategies to mitigate their impact in Senegal.

## Introduction

Arboviruses transmitted by mosquitoes pose a growing global health threat, significantly impacting morbidity and mortality. Among these, chikungunya (CHIKV) and o’nyong-nyong (ONNV) viruses are notable members of the alphavirus genus, recognized for their substantial effects on human health (1, 2). CHIKV, first isolated in Tanzania in 1952, has caused major outbreaks across Africa, Asia, and the Americas (3–5). Its transmission primarily involves *Aedes* mosquitoes, and its capacity for widespread dissemination was highlighted by the 2004 epidemic that originated in Kenya and rapidly extended to various regions, including the Indian Ocean islands and South and Southeast Asia (1, 5–7). By 2011, CHIKV reached the Western Hemisphere, spreading through the Caribbean and South America by 2014, resulting in hundreds of thousands of infections (8–10).

ONNV, identified in Uganda in 1959, has caused significant epidemics primarily in East and West Africa, with major outbreaks recorded in the 1950s and early 1990s (11–13). Transmitted by *Anopheles* mosquitoes, ONNV remains largely confined to the African continent (6). The first major epidemic occurred from 1959 to 1962, affecting over 2 million people across East and West Africa (14, 15). A subsequent outbreak from 1996 to 1997 primarily impacted Uganda, Kenya, and Tanzania, with infection rates ranging from 45-68% (14, 16). Despite its geographical confinement, ONNV has been detected in travelers returning from endemic regions (17).

Clinically, infections with CHIKV and ONNV are generally self-limiting but can present a spectrum of symptoms including fever, arthralgia, rash, headache, and malaise (18, 19). CHIKV is particularly associated with persistent arthralgia in patients (20). Although ONNV is endemic to Africa, comprehensive data on its morbidity and mortality are limited due to inadequate surveillance. The clinical features of alphavirus infections closely resemble those caused by other pathogens including *Plasmodium* (malaria) or *Salmonella* (typhoid fever) species, which are endemic in many West African countries (21–26). This symptom overlap complicates diagnosis and can lead to underreporting of CHIKV and ONNV, as their symptoms may be misattributed to other febrile illnesses.

In West Africa, including Senegal, autochthonous cases of CHIKV have been reported since 1960, with periodic outbreaks continuing up to 2023 (27–31). Despite these occurrences, current data on the seroprevalence of CHIKV and ONNV in Senegal remains limited. This study aims to address this knowledge gap by investigating the seroprevalence of CHIKV and ONNV in Senegal. Given the antigenic similarities between these viruses, our approach focuses on using microneutralization assays to help differentiate between them to provide a more in-depth understanding of their prevalence. By enhancing our knowledge of the epidemiological landscape of CHIKV and ONNV in Senegal, we hope to inform public health strategies and contribute valuable insights into the broader context of arboviral transmission in West Africa.

## Methods

### Study Population and Ethics Statement

The samples included in our study were originally collected as part of malaria and non-malarial surveillance among residents of Thies, Sindia, and Kedougou, Senegal. Samples from Thies and Sindia were part of the surveillance of non-malarial febrile illnesses, while samples from Kedougou were part of genomic surveillance of malaria. Informed consent was obtained from all subjects and/or their legal guardians for the initial sample collection as well as for its future use. All methods were performed in accordance with the guidelines and regulations set forth by the Declaration of Helsinki. The primary studies under which the samples and data were collected received ethical clearance from the CIGASS Institutional Review Board (IRB) (Protocol numbers: SEN15/46, 19300, and SEN14/49). All excess samples and corresponding data were banked and de-identified prior to the analyses. This study received and exemption determination from the Rutgers Robert Wood Johnson Medical School IRB.

### CHIKV viral like particle (VLP) ELISA

CHIKV VLPs (20 ng per well; Native Antigen) were coated onto Nunc Maxisorp 96-well plates (ThermoFisher) overnight, followed by blocking (1X PBST, 1% BSA; ThermoFisher) for 1 hour, then washing (1X PBST three times) and incubation with primary antibodies for 2 hours (serum at 1:400 dilution) at room temperature. The plates were washed three times with 1X PBST and bound sera were reacted with secondary antibodies (anti-human IgG conjugated with horseradish peroxidase [HRP]; ThermoFisher), according to the manufacturer’s instructions. The plates were washed three more times and allowed to develop with 1-Step TMB ELISA Substrate (ThermoFisher), according to the manufacturer’s instructions. The OD at 450 nm was read with a reference wavelength of 650 nm using an ELISA reader (Diasource). We interpreted the ELISA assay as follows: 1) sample absorbance / negative control absorbance ≤ 1.5, negative; 2) sample absorbance / negative control absorbance > 1.5, positive.

### Serum Microneutralization Assays

Microneutralization assays were performed to detect neutralizing antibodies against CHIKV and ONNV in samples that were IgG positive for CHIKV VLP. Sera were diluted in a two-fold series from 1:40 to 1:10,240 in PBS in a 96-well plate. For each dilution, contact was performed with 100 plaque forming units (PFU) of CHIKV (BEI Resources: NR-56523) or ONNV (BEI Resources: NR-51661) for one hour at 37°C in a 5% CO_2_ incubator. The complexes were then added to Vero cells (ATCC) CCL-81, 1.3 x 10^5^ cells per well. After 3-4 days of incubation, the cytopathogenic effects were investigated by 0.2% crystal violet (SigmaAldrich) staining. After staining for 2-4 hours, the plates were washed with copious amounts of tap water. For negative controls, at least 3 alphavirus-naïve serum samples were used to incubate with CHIKV or ONNV prior to pipetting onto the Vero cells per plate. For positive controls, 100 PFU of CHIKV or ONNV without any contact with serum were placed on Vero cells in triplicate per plate.

The neutralizing titer was defined as the inverse of the highest dilution resulting in an infectious reduction of 50%. A titer lower than 1:40 was considered as negative if cytopathogenic effects were not observed. Samples were considered as positive if the titer was ≥1:40, i.e., positive for ONNV if ONNV titers were at least two-fold higher than CHIKV titers, and CHIKV positive if CHIKV titers were at least four-fold higher than ONNV titers (32). The threshold was lower for ONNV than CHIKV because of the unique one-way cross-reactivity between CHIKV and ONNV, i.e., CHIKV antibodies are more likely to cross-react with ONNV antigens than ONNV antibodies with CHIKV antigens (33). All other results were interpreted as equivocal. 20 Senegalese serum samples that were previously determined to be alphavirus-naïve, were used to validate the assay. Each assay used at least 4 alphavirus-naïve serum samples as negative controls.

### Statistical Analysis

Categorical variables are represented as case counts and proportions, with differences assessed using the Chi-square or Fisher exact test as appropriate. To estimate the association between seroprevalence of alphavirus and demographics factors, including age group, gender, city, a univariate binary logistic regression analysis was performed. The adjusted odd ratios of the independent variables were computed by adjusting for age and gender as confounding variables. Statistical analyses were conducted using IBM SPSS statistics software version 28. The map of Senegal, heatmap, and graph representing rates and percentages of CHIKV and ONNV neutralizing antibodies were generated using R version 4.3.2 and Prism version 10.2.0.

## Results

### Characteristics of study subjects

A total of 470 samples collected from non-febrile subjects were included in the study. These samples were collected from three locations in Senegal: Sindia (n=94), Thies (n=94), and Kedougou (n=282). The samples from Sindia and Thies were malaria-negative by rapid diagnostic testing, whereas the samples from Kedougou were from subjects with malaria. The study population were relatively young, with approximately 48% of them aged 20 years or younger, and consisted of fewer females, comprising 44.89%. (Table 1).

**Table 1.** Baseline characteristics and chikungunya virus (CHIKV) and o’nyong-nyong virus (ONNV) serology.

|  | **Sindia**<br>**(n=94)** | **Thies**<br>**(n=94)** | **Kedougou**<br>**(n=282)** | **Total**<br>**(n=470)** |
| --- | --- | --- | --- | --- |
| **Demographics** |  |  |  |  |
| Year(s) samples collected | 2018 | 2018 | 2022-2023 |  |
| Age group, n (%) |  |  |  |  |
| ≤ 20 years | 40 (42.6) | 48 (41.1) | 136 (48.2) | 224 (47.7) |
| 21-40 years | 42 (44.7) | 23 (24.4) | 123 (43.6) | 188 (40.0) |
| > 40 years | 11 (11.7) | 23 (24.4) | 21 (7.5) | 55 (11.7) |
| Unreported | 1 (1.0) | — | 2 (0.7) | 3 (0.6) |
| Female gender, n (%) | 39 (41.5) | 51 (54.3) | 121 (42.9) | 211 (44.9) |
| History of malaria infection, n (%) | 0 (0) | 0 (0) | 282 (100) | 282 (60) |
| **Serology** |  |  |  |  |
| CHIKV VLP IgG+, n (%) | 14 (14.9) | 30 (31.9) | 137 (48.6) | 181 (38.5) |
| NT50, n (%) |  |  |  |  |
| CHIKV | 6 (6.4) | 18 (19.1) | 11 (3.9) | 35 (7.4) |
| ONNV | 4 (4.3) | 4 (4.3) | 38 (13.5) | 46 (9.8) |
| Equivocal | 4 (4.3) | 5 (5.3) | 15 (5.3) | 24 (5.1) |
| Negative | 0 (0.0) | 0 (0.0) | 12 (4.3) | 12 (2.6) |

#### Prevalence of CHIKV and ONNV

We first employed an anti-CHIKV VLP IgG ELISA to screen 470 non-febrile serum samples. The CHIKV VLP antigen is specifically designed to detect IgG antibodies against CHIKV but can also recognize cross-reactive antibodies against related alphaviruses. Of the 470 samples tested, 181 were positive, resulting in an estimated alphavirus prevalence of 38.5%. Seroprevalence rates varied by location. Sindia had a rate of 14.9%, Thies had 31.9%, and Kedougou had 48.6% (Table 1).

Moreover, the microneutralization assay, regarded as the gold standard for identifying prior exposure to a specific virus was utilized to differentiate exposure to CHIKV and/or ONNV. Among the CHIKV IgG positive samples, 117 were available to assess neutralizing antibodies against CHIKV and/or ONNV. The prevalence of neutralizing antibodies against CHIKV and ONNV varied by location, gender, and age group. The prevalence rates of CHIKV neutralizing antibodies were 6.4% in Sindia, 19.1% in Thies, and 3.9% in Kedougou (Table 1). By gender, 7.0% for males and 8.0% for females had CHIKV neutralizing antibodies (Table 2). Across age groups, the prevalence was 7.6% in subjects aged ≤20 years, 5.3% in subjects aged 21 to 40 years, and 14.8% in those over 40 years (Table 2). For ONNV, the neutralizing antibody prevalence rates were 4.3% in Sindia, 4.3% in Thies, and 13.5% in Kedougou (Table 1). The prevalence of ONNV neutralizing antibodies was 10.2% for males and 9.5% for females (Table 2). Age-stratified prevalence showed a rate of 8.5 % in subjects aged ≤20 years, 8.0% in subjects aged 21 to 40 years, and 20% in those over 40 years (Table 2).

**Table 2.** Baseline characteristics of NT50-confirmed chikungunya virus (CHIKV) and o’nyong-nyong virus (ONNV) subjects.

|  | **Total**<br>**n= 470** | **CHIKV NT50-**<br>**confirmed subjects**<br>**n=35** | **ONNV NT50-**<br>**confirmed subjects**<br>**n=46** |
| --- | --- | --- | --- |
| Age group, n (%) |  |  |  |
| ≤ 20 years | 224 (47.7) | 17 (7.6) | 19 (8.5) |
| 21-40 years | 188 (40.0) | 10 (5.3) | 15 (8.0) |
| > 40 years | 55 (11.7) | 8 (14.8) | 11 (20.0) |
| Unreported | 3 (0.6) | 0 (0.0) | 1 (33.3) |
| Gender, n (%) |  |  |  |
| Male | 256 (54.5) | 18 (7.0) | 26 (10.2) |
| Female | 211 (44.9) | 17 (8.0) | 20 (9.5) |
| Unreported | 3 (0.6) | 0 (0.0) | 0 (0.0) |
| City, n (%) |  |  |  |
| Sindia | 94 (20.0) | 6 (6.4) | 4 (4.3) |
| Thies | 94 (20.0) | 18 (19.1) | 4 (4.3) |
| Kedougou | 282 (60.0) | 11 (3.9) | 38 (13.5) |
| History of<br>malaria<br>infection, n (%) | 282 (60) | 11 (3.9) | 38 (13.5) |

Overall, 7.4% of the study subjects had neutralizing antibodies against CHIKV, 9.8% against ONNV, 5.1% were equivocal for both CHIKV and ONNV, and 2.6% were negative, indicating prior exposure to other related alphaviruses (Table 1, Fig. 2A). Moreover, of the subjects with CHIKV neutralizing antibodies, 82.9% had cross-neutralizing antibodies against ONNV (Fig. 1B-C). However, subjects with ONNV neutralizing antibodies, 71.7% had cross-neutralizing antibodies against CHIKV (Fig. 2B-C).

**Fig 1.**
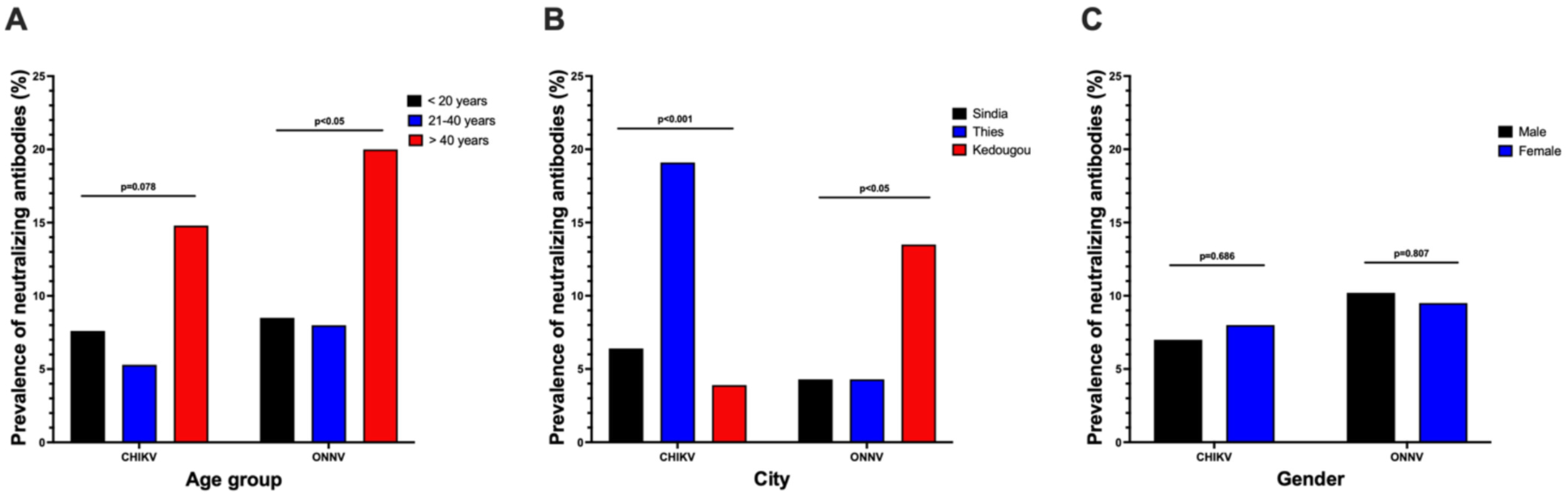
Comparison of the prevalence of neutralizing antibodies to CHIKV and ONNV by age group, city, and gender. Differences between (A) age, (B) city, and (C) gender were assessed using the Chi-square test or Fisher’s exact test, as appropriate. P-values less than 0.05 were considered to be statistically significant, while values less than 0.1 were considered to be marginally significant.

**Fig. 2.**
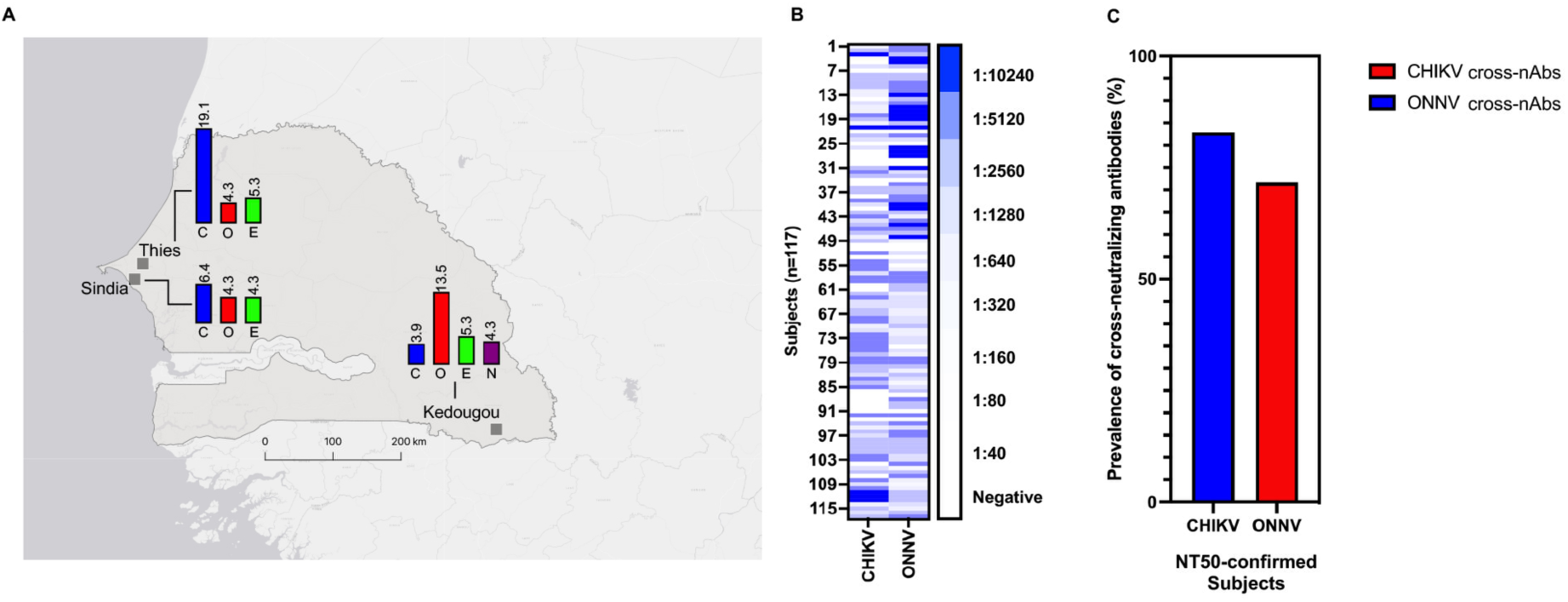
Antibody neutralization against chikungunya (CHIKV) and o’nyong-nyong (ONNV) viruses in Senegal. (A) Map depicts locations in Senegal (Sindia, Thies, and Kedougou) where samples were collected from non-febrile subjects. For each location, bar charts represent the neutralizing antibody prevalence against CHIKV (C) or ONNV (C), or equivocal (E) and negative (N) results. Prevalence values are listed above each bar. (B) CHIKV virus-like particle IgG-positive samples (n=117) were tested for neutralizing antibodies against CHIKV and ONNV. Colors depict the 50% neutralizing antibody titers (NT50), each row represents one individual, and each column is the titer to CHIKV or ONNV. (C) The percentage of NT50-confirmed CHIKV or ONNV subjects with cross-neutralizing antibodies to the heterologous virus.

#### Risk factors for CHIKV and ONNV infection

Univariate logistic regression, adjusted for age and gender, revealed that being a resident of Thies (aOR, 2.384; 95% CI: 1.151-4.939) or Kedougou (aOR, 5.591; 95% CI: 3.001-10.418) were associated with increased odds of exposure to an alphavirus based on IgG reactivity to CHIKV VLP, with the highest risk observed in Kedougou (S1 Table). Modeling the presence of neutralizing antibodies to CHIKV against background factors showed that subjects aged over 40 years had higher odds of CHIKV infection after adjusting for age and gender (aOR, 2.094; 95% CI: 0.846-5.185), though this association was marginally significant (Table 3). Additionally, the prevalence of CHIKV neutralizing antibodies was higher in subjects aged over 40 years compared to the other age groups (Fig 1A). Furthermore, subjects from Thies were more likely to be infected with CHIKV (aOR, 3.147; 95% CI: 1.164-8.510) (Table 3), with a significantly higher prevalence of CHIKV neutralizing antibodies in this city compared to Sindia and Kedougou (Fig 1B). Residence of Kedougou was associated with significantly lower odds of contracting CHIKV (aOR, 0.606; 95% CI: 0.217-1.692) (Table 3).

**Table 3.**
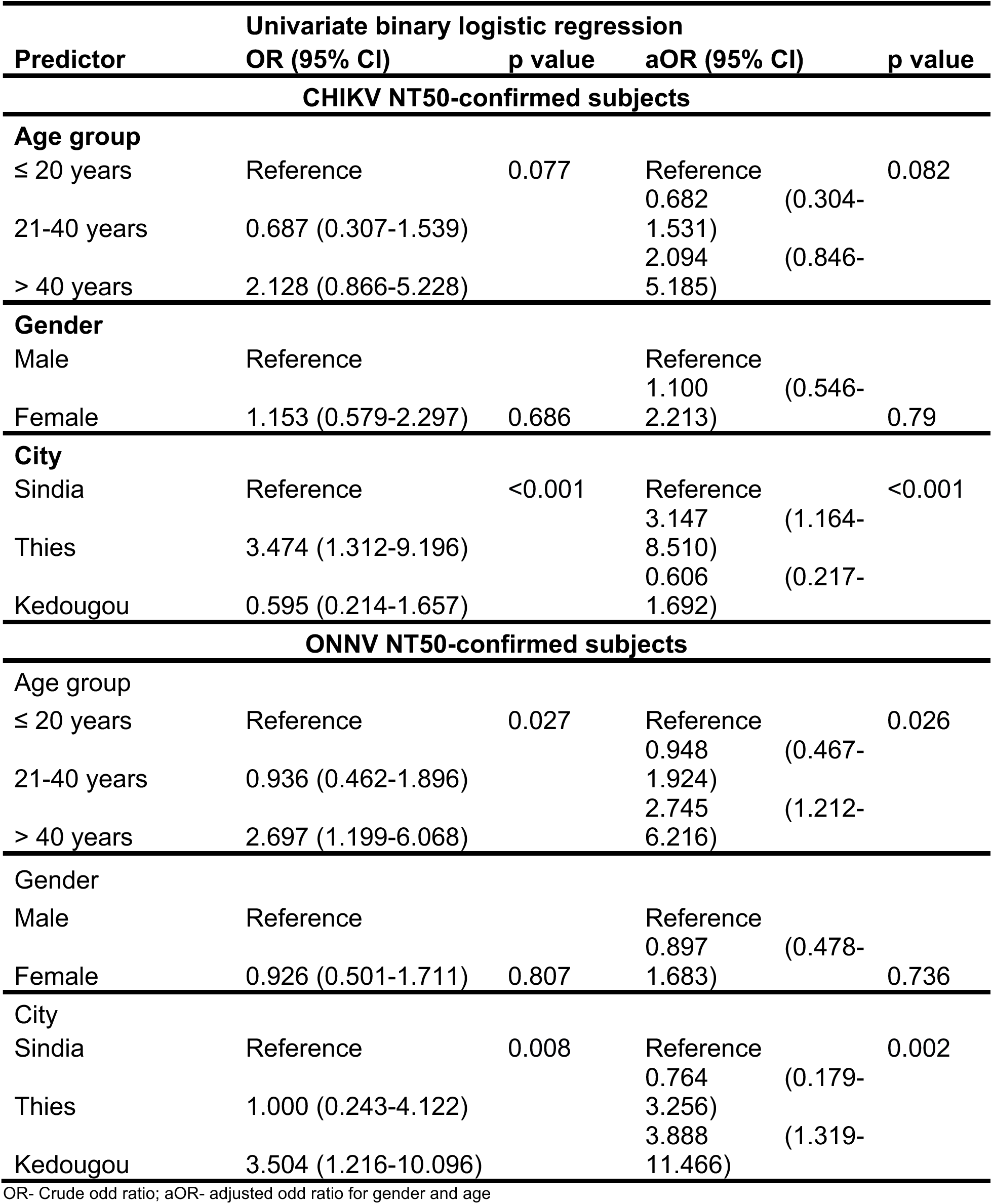
Risk factors for chikungunya virus (CHIKV) and o’nyong-nyong virus (ONNV) infections in Senegal.

| Predictor | Univariate binary logistic regression |  |  |  |
| --- | --- | --- | --- | --- |
|  | OR (95% CI) | p value | aOR (95% CI) | p value |
| **CHIKV NT50-confirmed subjects** |  |  |  |  |
| **Age group** |  |  |  |  |
| ≤ 20 years | Reference | 0.077 | Reference | 0.082 |
| 21-40 years | 0.687 (0.307-1.539) |  | 0.682 (0.304-1.531) |  |
| > 40 years | 2.128 (0.866-5.228) |  | 2.094 (0.846-5.185) |  |
| **Gender** |  |  |  |  |
| Male | Reference |  | Reference |  |
| Female | 1.153 (0.579-2.297) | 0.686 | 1.100 (0.546-2.213) | 0.79 |
| **City** |  |  |  |  |
| Sindia | Reference | <0.001 | Reference | <0.001 |
| Thies | 3.474 (1.312-9.196) |  | 3.147 (1.164-8.510) |  |
| Kedougou | 0.595 (0.214-1.657) |  | 0.606 (0.217-1.692) |  |
| **ONNV NT50-confirmed subjects** |  |  |  |  |
| **Age group** |  |  |  |  |
| ≤ 20 years | Reference | 0.027 | Reference | 0.026 |
| 21-40 years | 0.936 (0.462-1.896) |  | 0.948 (0.467-1.924) |  |
| > 40 years | 2.697 (1.199-6.068) |  | 2.745 (1.212-6.216) |  |
| **Gender** |  |  |  |  |
| Male | Reference |  | Reference |  |
| Female | 0.926 (0.501-1.711) | 0.807 | 0.897 (0.478-1.683) | 0.736 |
| **City** |  |  |  |  |
| Sindia | Reference | 0.008 | Reference | 0.002 |
| Thies | 1.000 (0.243-4.122) |  | 0.764 (0.179-3.256) |  |
| Kedougou | 3.504 (1.216-10.096) |  | 3.888 (1.319-11.466) |  |

OR- Crude odd ratio; aOR- adjusted odd ratio for gender and age

For ONNV, univariate analysis, adjusted for age and gender, revealed that subjects aged over 40 years (aOR, 2.745; 95% CI: 1.212-6.216) had significantly increased odds of infection (Table 3), with a significantly higher prevalence of ONVV neutralizing antibodies in this age group compared to the other age groups (Fig 1A). Subjects aged 21 to 40 (aOR, 0.682; 95% CI: 0.304-1.531) years were associated with lower odds of ONNV infection (Table 3). Being a resident of Kedougou (aOR, 3.888; 95% CI: 1.319-11.466) was associated with increase odds of ONNV infection (Table 3), with significantly higher prevalence of ONVV neutralizing antibodies compared to the other cities (Fig 2B). Residence from Thies was associated with lower odds of infection (aOR, 0.764; 95% CI: 0.179-3.256) (Table 3).

## Discussion

In this study, we observed an overall alphavirus prevalence of 38.5% with notable geographical variations. Virus-specific analysis revealed prevalence rates of 7.4% for CHIKV exposure and 9.8% for ONNV, both exhibiting significant geographical and age-related variations. Cross-reactivity analysis showed that 82.9% of CHIKV cases exhibited a neutralizing response to ONNV, while 71.7% of ONNV cases cross-neutralized CHIKV. Older age emerged as a significant risk factor for ONNV exposure, and to a lesser extent CHIKV. Additionally, ONNV exposure was significantly associated with residence in Kedougou, while CHIKV exposure was strongly linked to residence in Thies. These findings emphasize the continued transmission of alphaviruses in Senegal, specifically CHIKV and ONNV, driven by geographic location as well as age-susceptibility.

The first documentation of a CHIKV human infection dates back to 1966 in Rufisque, a city near Dakar, as part of arbovirus surveillance programs led by the Institut Pasteur de Dakar. During this surveillance, CHIKV was detected in both mosquito vectors and in human populations (29). Since then, numerous reports have documented CHIKV circulation in the country, highlighting its endemicity and periodic re-emergence. A notable example is the recent outbreak of CHIKV in Kedougou, which was attributed to the re-emergence of an endemic strain (27).

To date, seroprevalence studies aimed at understanding CHIKV circulation in Senegal have predominantly relied on detecting anti-CHIKV IgG antibodies using ELISA (34, 35). However, this approach tends to overestimate CHIKV prevalence due to the significant cross-reactivity between CHIKV and related alphaviruses, particularly ONNV. This limitation highlights the need for more specific diagnostic methods, such as virus neutralization assays or molecular techniques, to accurately discern CHIKV exposure and better differentiate it from other co-circulating alphaviruses. Consequently, the use of a microneutralization assay in this study allowed us to identify subjects with specific or cross-reactive responses to either CHIKV or ONNV, providing a more accurate assessment of their respective seroprevalence rates.

The prevalence rates for CHIKV and ONNV observed in our study differs from those reported in similar studies in Senegal and other countries. For instance, a recent study in Kedougou, involving inhabitants of 120 randomly selected villages, reported a CHIKV prevalence of 54% (35), which is higher than both our overall observed rate and the prevalence among the subjects from Kedougou in our study. An explanation for the higher rate in this study is likely due to implementation of an anti-CHIKV IgG ELISA, which measures cross-reactive alphavirus antibodies. In Nigeria, a systematic review encompassing published literature spanning four decades reported a CHIKV prevalence of 46.1% (36) based on neutralizing antibodies, a rate higher than that observed in the present study. Additionally, our observed CHIKV prevalence rate is lower than the 56.5% (37) reported among blood donors in Burkina Faso. Conversely, a study conducted in Kenya reported CHIKV prevalence of 9.8% (38) among children from four communities within the coastal and western regions, a rate comparable to our findings.

For ONNV, the prevalence rate observed in our study is lower than the 54.5% (37) reported among blood donors in Burkina Faso, and the 28.4% (39) reported among HIV patients in Madagascar. Importantly, both studies used anti-CHIKV IgG ELISAs to measure antibody responses, which may have skewed the higher rates. Additionally, modeling studies in Mali, using serological data estimated an ONNV prevalence of 30% (33), also exceeding our observed rate. These discrepancies may be attributed to differences in study population, sample size, methodological approaches, and ecological or environment factors that influence virus transmission.

Our univariate logistic regression analysis identified older age as a risk factor for both CHIKV and ONNV infection. We observed a higher prevalence of neutralizing antibodies for both viruses in the older age group compared to their younger counterparts, corroborating the notion that cumulative exposure over time contributes to higher exposure rates. This demographic insight is crucial for developing targeted public health strategies, especially for vaccine deployment and vector control initiatives. The identification of older age as a risk factor for CHIKV infection is consistent with similar findings from a study in Burkina Faso (37).

Furthermore, residents of Thies showed a three-fold increase in the likelihood of CHIKV infection compared to other regions, as evidenced by the higher prevalence of CHIKV neutralizing antibodies in Thies. Conversely, residents of Kedougou showed a four-fold increase in the likelihood of ONNV infection, as evidenced by higher prevalence of ONNV neutralizing antibodies compared to the other regions. These regional differences may be attributed to distinct local vector ecologies or differences in human activities that increase exposure to virus-specific mosquito vectors. These hypotheses require further research.

We also observed that the prevalence of cross-neutralizing antibodies against ONNV was higher among subjects with CHIKV neutralizing antibodies compared to those with ONNV neutralizing antibodies exhibiting cross-neutralization against CHIKV. These findings corroborate a directional bias in immune cross-reactivity, where CHIKV exposure might confer broader immunity, potentially impacting subsequent ONNV infection dynamics (33). This asymmetric cross-reactivity could also be due to structural similarities in the viral envelope proteins of CHIKV and ONNV, with CHIKV eliciting a broader neutralizing response (33). This phenomenon is crucial for diagnostic tests, which may need differentiation between past infections of these two viruses based on neutralizing antibody profiles. Moreover, understanding the mechanisms behind this differential cross-reactivity could guide vaccine strategies where a vaccine against one virus might offer partial protection against the other, hence optimizing the control measures for both viruses in regions where they co-circulate.

One of the strengths of this study is our robust delineation of subjects previously exposed to CHIKV or ONNV in Senegal, achieved through the use of neutralization assays to minimize cross-reactivity. This methodological rigor makes our study the first to report the co-circulation of these antigenically related viruses in the country, providing valuable insights into their epidemiology and the public health risks they pose.

Despite its strengths, our study has several limitations. Of the 180 samples positive for alphavirus IgG by ELISA, we were only able to perform neutralization assays on 117 due to sample availability. This limitation likely led to an underestimation of the true prevalence of CHIKV and ONNV in our study populations. Additionally, while our use of neutralization assays minimized cross-reactivity compared to the ELISA, a large percentage of samples were considered equivocal, consistent with findings from other studies. In contrast, other studies that rely solely on ELISA often classify these equivocal results as positive, leading to potential overestimations of CHIKV prevalence (34, 35, 37). Our approach provides a more conservative and potentially accurate estimate but underscores the need for improved diagnostic tools to address the challenge of equivocal results in alphavirus seroprevalence studies. Moreover, without concurrent clinical data, we also cannot definitively link our serological findings to symptomatic disease cases, underestimating the true burden of these infections. Additionally, the insights into antibody cross-reactivity are essential for the interpretation of seroprevalence data in regions endemic to both viruses, as they could lead to overestimations of immunity against one virus due to past exposure to the other. This is particularly relevant in vaccine efficacy trials and in the evaluation of natural immunity in the population.

Conclusions drawn from our research illustrate the persistent circulation of CHIKV and ONNV in Senegal, necessitating sustained surveillance and public health preparedness to mitigate the impact of these and other arboviruses. The findings also emphasize the importance of ecological and demographic factors in shaping the epidemiology of these diseases, highlighting the need for comprehensive arbovirus control strategies that integrate vector management with community health initiatives.

Future studies should aim to incorporate entomological and clinical data to provide a more holistic understanding of the transmission dynamics and to facilitate the development of targeted interventions, including vaccines and therapeutic measures.

## Data Availability

Data produced as part of the current study are available in the manuscript or upon request from the corresponding author.

## Acknowledgements

We would like to thank Rutgers Global Health Institute, Rutgers Robert Wood Johnson Medical School, and the Child Health Institute of New Jersey.

## Author Contributions

Conceptualization: BBH

Data curation: PBT, MS, IMN, MT, AG, MA, BBH, DN

Formal analysis: PBT, MS, IMN, MT, AG, MA, BBH, DN

Funding acquisition: BBH, DN

Investigation: PBT, MS, IMN, MT, AG, MA, BBH, DN

Methodology: PBT, MS, IMN, MT, AG, MA, AMM, AKD, MAD, JFG, MSY, YD, BD, KD, MCS, ASB, BBH, DN

Project administration: BBH, DN

Resources: BBH, DN

Software: PBT, MS, BBH

Supervision: BBH, MS, DN

Validation: PBT, MS, IMN, MT, AG, MA, BBH, DN

Visualization: PBT, MS, IMN, MT, AG, MA, BBH, DN

Writing – original draft: PBT, BBH

Writing – review & editing: PBT, MS, IMN, MT, AG, MA, AMM, AKD, MAD, JFG, MSY, YD, BD, KD, MCS, ASB, BBH, DN

